# New job, new habits? A Multilevel Interrupted Time Series analysis of diet, physical activity and sleep changes among young adults starting work for the first time

**DOI:** 10.1101/2024.07.17.24310567

**Authors:** Alena F Oxenham, Tanya Braune, Esther van Sluijs, Hannah Fairbrother, Adam Martin, Eleanor M Winpenny

## Abstract

**Background:** The workplace is an important determinant of health that people are exposed to for the first-time during adolescence/early adulthood. This study investigates how diet, physical activity, and sleep change as people aged 16-30 years transition into work and whether this varies for different individuals and job types.

**Methods:** Multilevel linear regression models assessed changes in fruit and vegetable intake, sleep duration, and physical activity among 3,302 UK Household Longitudinal Study (UKHLS) participants aged 16-30 years, who started work for the first time between 2015 and 2023. In line with interrupted time series, models assessed behavioural trends in the period before starting work, the immediate effect of starting work, and changes in behaviour over time after employment. Stratified analyses examined differences by selected individual and job characteristics, adjusted for covariates. All analyses were conducted in R v.4.3.2.

**Results:** Sleep duration was stable over the years before and after starting work, but starting work was associated with an immediate reduction in sleep duration (*β*=-9.74 [95% CI: −16.81 to −2.67], min/night). Physical activity, measured in Metabolic Equivalent Tasks (MET), increased immediately after starting work (*β =* 113.3, [95% CI: 80.49 to 146.11] MET-min/day), but subsequently decreased over time after starting work (*β =* −26.7, [95% CI: −40.75 to −12.66] MET-min/day/year). The increase in physical activity was greater among men, among those with no degree and among those starting lower socioeconomic classification jobs. Starting a “work from home” job had an immediate negative effect on physical activity (*β =* −157.81, [-313.07 to - 2.56] MET-min/day), whereas those who worked at their employer’s premises showed an initial increase (*β=* 125.71 [95% CI: 85.79 to 165.63] MET-min/day). Starting work had little influence of fruit and vegetable consumption.

**Conclusions:** This is the first study to examine how diet, physical activity, and sleep behaviours in young adults change as they start employment in the UK. Starting work is associated with decreased sleep time and increased physical activity, with differences based on sociodemographic and job characteristics. Future research should consider these potential influences of the work environment when developing intervention targets to promote healthy behaviour in the workplace.

## Introduction

Healthy lifestyles, including behaviours such as adequate sleep, regular physical activity, and a balanced diet, are fundamental to long-term well-being and good physical and mental health (1). Young adulthood, defined here as ages 16 to 30 is a critical period in health development. Whilst this is typically a period of peak physical health, young adults may also be developing risk factors for chronic disease that arise later in life (2–5). Notably, obesity prevalence rises sharply during young adulthood, with the majority of the United Kingdom (UK) population living with overweight or obesity by their early thirties (6). This transitional phase can also involves significant life changes, including transitioning from education to employment, and from familial homes to independent living, all of which might have a detrimental effect on health behaviours including diet, physical activity, and sleep patterns (7). Understanding the unique challenges and opportunities of the transition from adolescence to young adulthood for the development of long-term health behaviour habits is crucial to supporting healthy adult lifestyles (2,8,9).

Starting work is one key transition of young adulthood, encompassing changes in physical and social environments, daily routines and activities, and resources such as time and money, all of which are determinants of health behaviours and later life health (10). Transitions into employment are becoming increasingly more complex, as young people are spending more time in education, have longer spells of unemployment and spend more time working in part-time jobs (11). The employment conditions experienced by those starting work are also rapidly changing, with rises in remote and hybrid working since the COVID-19 pandemic, and increases in precarious employment and zero-hour contracts, all of which have impacts on health behaviours (12–14).

A number of previous studies have investigated changes in health behaviours on starting employment, but no studies have been conducted in the UK context. A recent systematic review on changes in physical activity and diet through young adulthood transitions found that two out of three longitudinal studies reported decreases in physical activity through the transition into employment, and one reported no change (15). Another scoping review reported that entering employment was associated with decreases in physical activity across several studies, although this was mostly in leisure time physical activity. Four studies in the review did not find any changes in physical activity, and one study reported differences based on sex such that men were less likely to show decreases in physical activity after starting work. (16). The Project Eating and Activity across Time (EAT) study, based in the United States, followed up participants aged 11-18 at baseline over four timepoints across fifteen years and assessed life events as well as moderate and vigorous activity. The authors reported that starting work in a full-time job (more than 30 hours) was not associated with any changes in physical activity (17). Overall, the current literature depicts mixed results of decreases or no changes in physical activity after starting work.

Studies examining changes in diet across the transition into employment have also found mixed results. A systematic review conducted in 2020 included only one study which examined entering employment and changes in diet, which reported no significant change after adjusting for confounders (15). A recent latent growth analysis using Project EAT data found that starting full-time work was associated with increases in diet quality, especially among men (18). Meanwhile, another analysis of Project EAT data found that starting work was associated with an increase in fast-food intake (19).

To our knowledge, the effect of starting work on sleep behaviours in young adults has not been studied, but previous longitudinal studies using cohort data from Australia and Brazil have found that a majority of young adults have a negative trajectory of sleep duration and sleep disturbances through the transition into young adulthood from adolescence (20,21).

Previous studies in adult populations have demonstrated the workplace as an important determinant of health (22–26), with factors such as working shifts, and irregular or long hours having a negative impact on sleep, diet, and physical activity (27–31). However, few studies have focused on young adults. One cross-sectional study showed that the healthfulness of the work food- and physical activity-environment (i.e. access to fast-food, sugar-sweetened beverages, gyms, walkability etc.) was positively associated with diet and physical activity levels in young adults (32). However, changes in health behaviours across the transition into the workplace are likely to depend on the characteristics of the job itself.

This study responds to the lack of evidence exploring changes in health behaviours on starting employment in the UK context. We examine the longitudinal relationship between entering employment and changes in young adults’ health behaviours, and how these differ by individual and job characteristics. Looking at individual-level change in health behaviours over time, and in response to the transition of starting work, provides a more causal understanding of how work and work-related factors may be contributing to the development of young adult health behaviour patterns. The aim of this paper is therefore to investigate how starting work for the first time is associated with changes in health behaviours (physical activity, diet and sleep duration), and explore the role of individual (sex, education level and parental social economic position) and job characteristics (shift times, commute mode, job location, work hours) in moderating this relationship.

## Methods

### Study Overview and Data Collection Participants and procedures

The UK Household Longitudinal Study (UKHLS) is a panel survey that has collected data annually since 2009 on approximately 40,000 UK households (33), with the most recent data available (wave 13) collected in 2022/2023. Data on all four health behaviours; physical activity, sleep, vegetable consumption, and fruit consumption, , were first consistently collected in the adult surveys from wave 7 onwards (2015–2017).

We included in these analyses UKHLS participants who started work between the ages of 16 and 30, and responded at least once before and once after starting work to the health behaviour questions in waves 7-13.

### Exposure variables

#### Starting Work

Participants were asked to identify their main occupation, from a list including “Self-employed”, “Paid Employment”, “Unemployed”, “On maternity leave”, “Family care or home”, “Full-time student”, “LT sick or disabled”, “Government training scheme”, “Unpaid, family business”, “On apprenticeship”, “On furlough”, “Temporarily laid off/short term working”, “On shared parental leave”, and “Doing something else”. Those identifying as working self-employed, full-time or part-time, or as an apprentice were defined as ‘working’. Those who reported that their main occupation was ‘education’ were not defined as ‘working’ as working was not their main occupation. A binary variable was generated indicating whether or not the participant had yet started work, coded as 0 for all waves of data collected before starting work and 1 for all waves of data collected after starting work (X_ti_), where starting work was defined as the participant first reporting working self-employed, full-time, part-time, or as an apprentice as their main occupation.

### Time relative to starting work

Using information on survey completion date (MM/YY) which is reported in UKHLS at every wave for all participants, a continuous variable was generated measuring time (years) in relation to the recorded date of the first wave of data collection after starting work (Z_ti_), such that Z_ti_<0 when X_ti_=0 and Z_ti_>=0 when X_ti_=1. In a sensitivity analysis, this variable (Z_ti_) was altered to instead measure time (months) since the reported job start date (MM/YY), a variable which is only available in UKHLS for a subsample of participants (see Figure 1).

**Figure 1:**
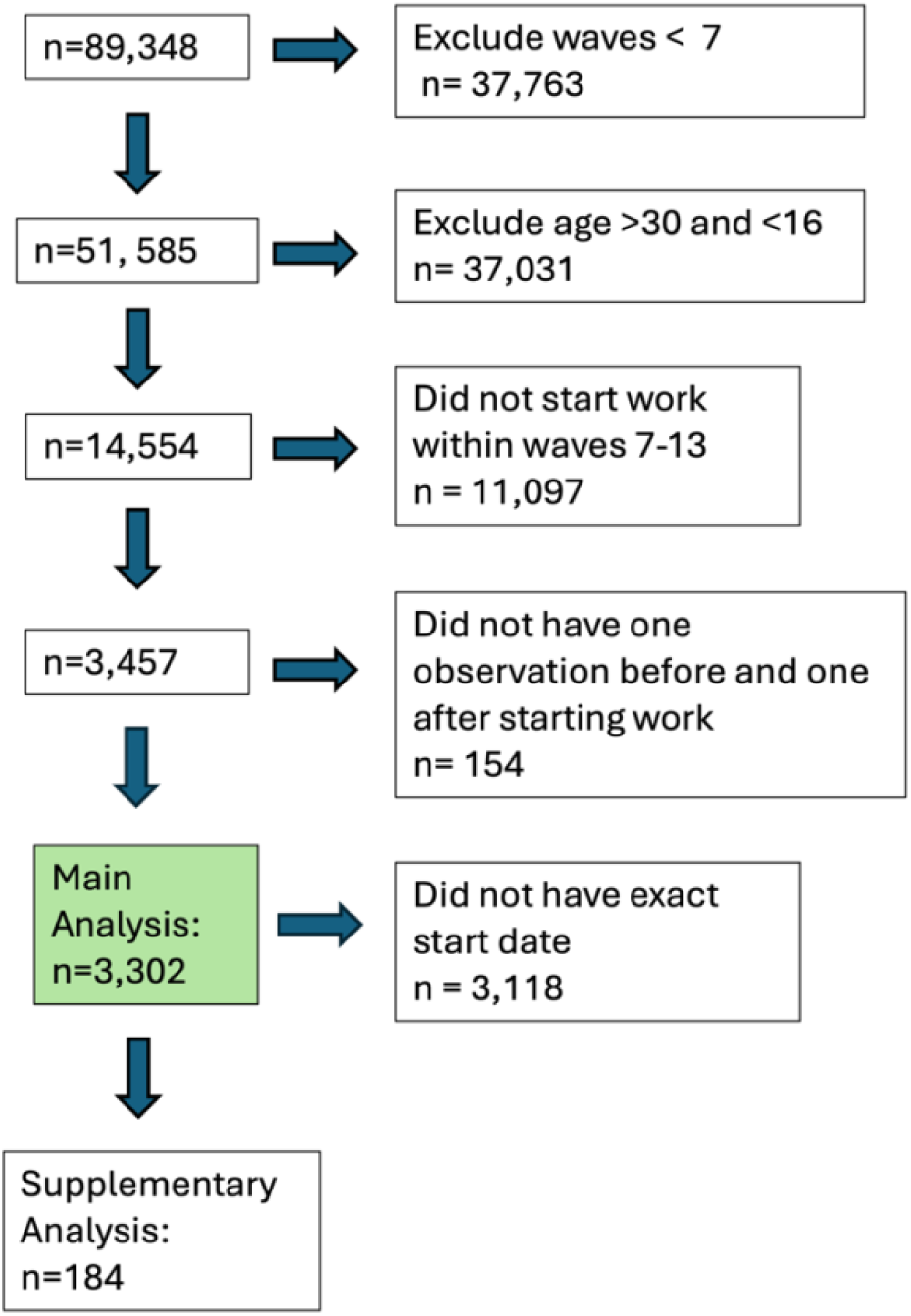
Inclusion Criteria Flowchart

### Outcome Variables (Y_ti_)

#### Diet

Vegetable and fruit consumption were measured in two separate questions for each participant every other wave (7,9,11, and 13) as shown in *Table 1.* The questions were adapted from HILDA and the Development of the Eating Choices Index (ECI) (32).

**Table 1.** Fruit and Vegetable questions and response options.

|  | Question | Response Options |
| --- | --- | --- |
| Fruit frequency | “Including tinned, frozen, dried and fresh fruit, on how many days in a usual week do you eat fruit?” | Never [coded as 0]<br>1-3 Days [2]<br>4-6 Days [5]<br>Every day [7] |
| Fruit amount | “On the days when you eat fruit, how many portions (e.g. an apple, an orange, some grapes) do you eat?” | Values $\geq 0$ |
| Vegetable frequency | “Including tinned, frozen and fresh vegetables, on how many days in a usual week do you eat vegetables? Do not include potatoes, crisps or chips.” | Never [coded as 0]<br>1-3 Days [2]<br>4-6 Days [5]<br>Every day [7] |
| Vegetable amount | “On the days when you eat vegetables, how many portions (i.e. 3 heaped tablespoons) do you eat? Please do not include potatoes.” | Values $\geq 0$ |

A daily vegetable portion measure was calculated by multiplying responses to vegetable frequency and vegetable portion questions and dividing by 7. Daily fruit consumption was assessed in a similar manner.

#### Sleep

Participants were asked every three waves (7,10,13): “How many hours of actual sleep did you usually get per night during the last month?" They were advised to indicate the most accurate reply for the majority of days and nights and that this may differ from the actual number of hours spent in bed.

### Physical Activity

Three variables from the validated International Physical Activity Questionnaire (IPAQ) (34) were used to assess participants’ weekly physical activity habits in waves 7, 9, 11, 12, and 13: moderate, vigorous and walking activity.

These responses were combined using the formula provided by the IPAQ guidelines to generate a weekly Metabolic Equivalent Tasks (MET)-minutes/week score (35). The final MET-minutes/week variable was then used to generate a daily MET-min/day estimate.

#### Moderators

Data on individual and job characteristics were self-reported at all survey waves. We used data reported at the first wave of data collection after participants started work. Data were recoded to give the following individual characteristics, with job characteristics presented in Table 2.

**Table 2:**
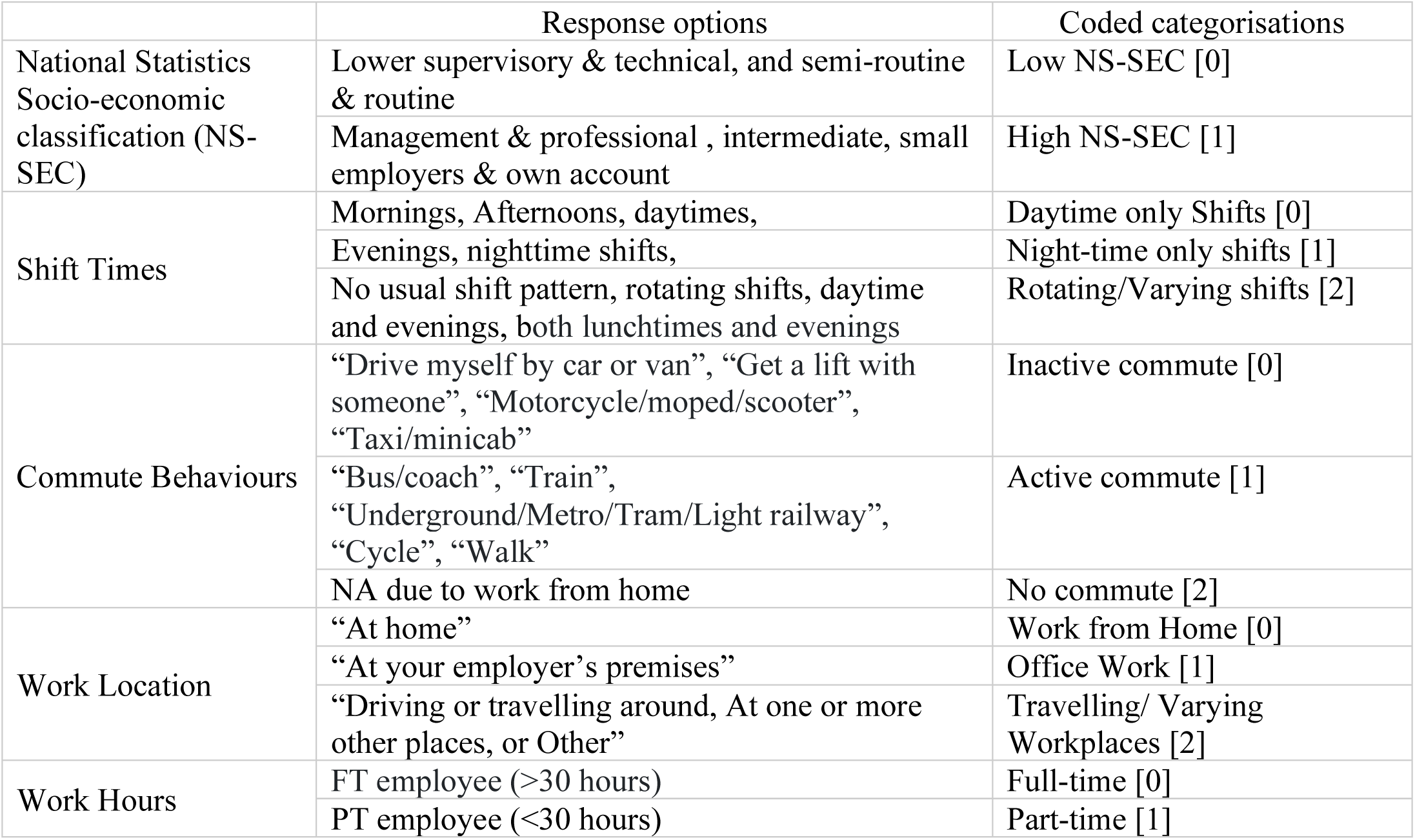
Job characteristics response options and categorisations.

**Table 3.**
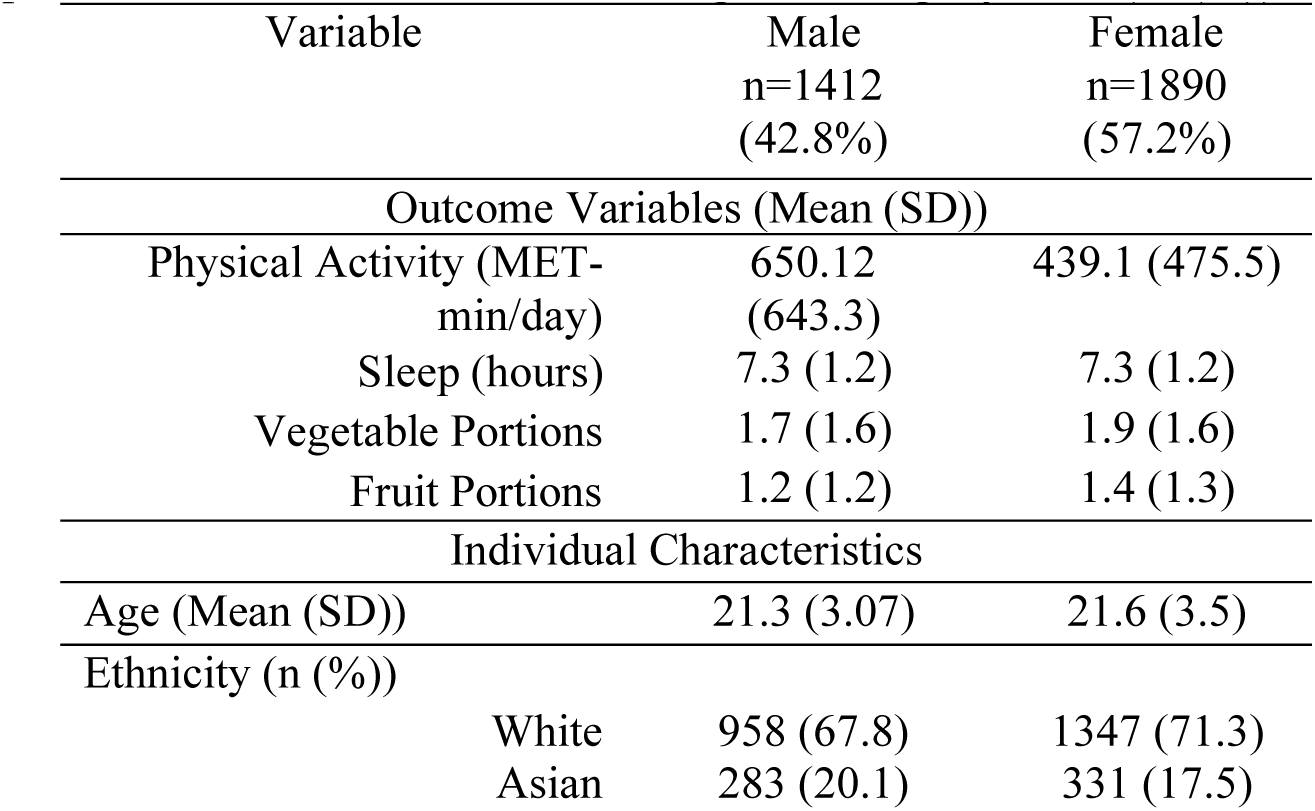
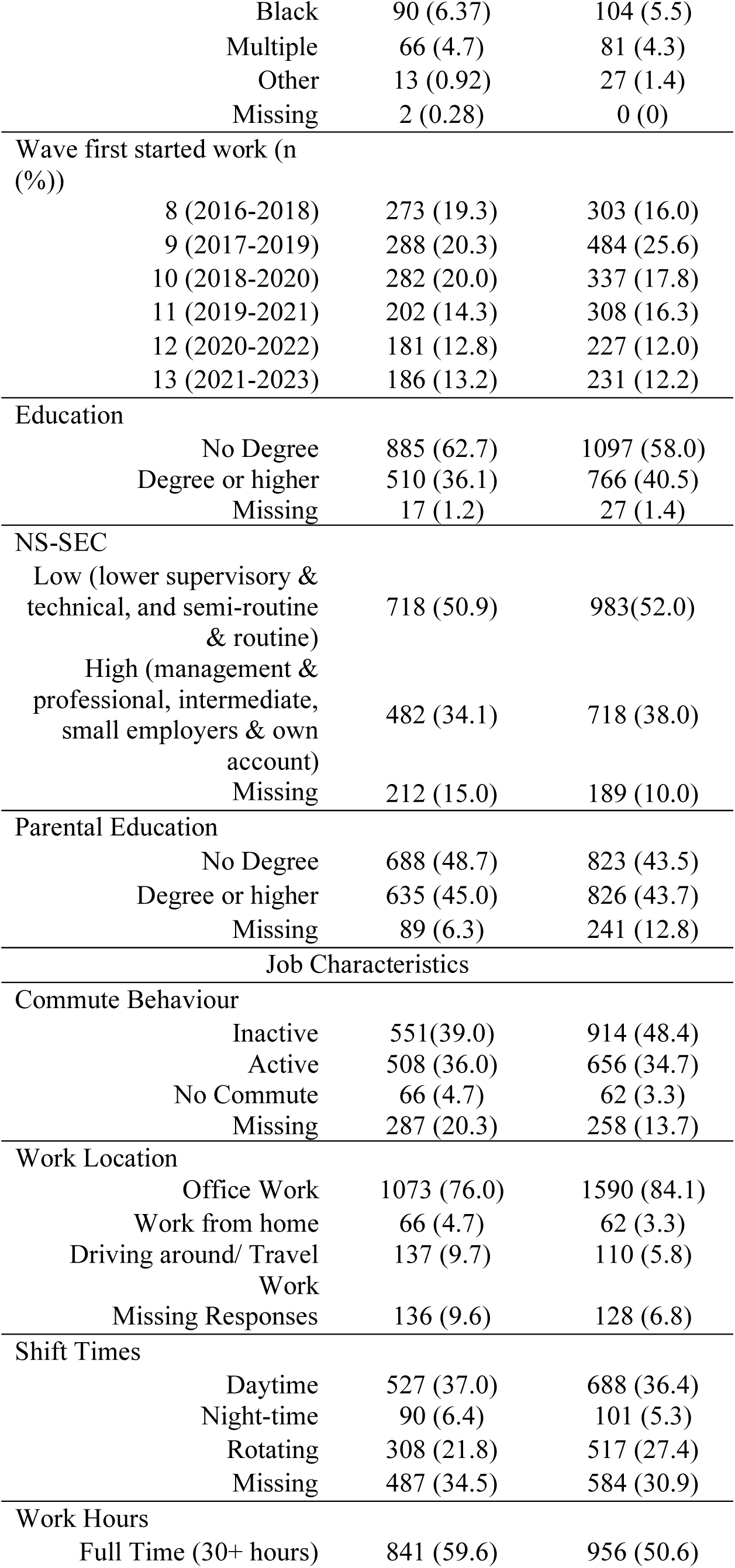
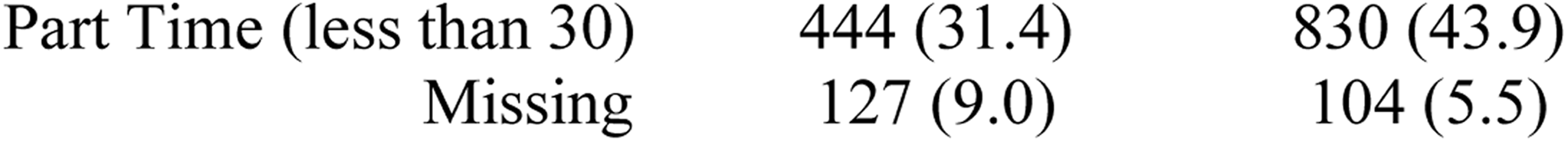
*Descriptive statistics* at first instance of reported employment (n; (%))

| Variable | Male<br>n=1412<br>(42.8%) | Female<br>n=1890<br>(57.2%) |
| --- | --- | --- |
| Outcome Variables (Mean (SD)) |  |  |
| Physical Activity (MET-min/day) | 650.12<br>(643.3) | 439.1 (475.5) |
| Sleep (hours) | 7.3 (1.2) | 7.3 (1.2) |
| Vegetable Portions | 1.7 (1.6) | 1.9 (1.6) |
| Fruit Portions | 1.2 (1.2) | 1.4 (1.3) |
| Individual Characteristics |  |  |
| Age (Mean (SD)) | 21.3 (3.07) | 21.6 (3.5) |
| Ethnicity (n (%)) |  |  |
| White | 958 (67.8) | 1347 (71.3) |
| Asian | 283 (20.1) | 331 (17.5) |

|  |  |  |  |
| --- | --- | --- | --- |
|  | Black | 90 (6.37) | 104 (5.5) |
|  | Multiple | 66 (4.7) | 81 (4.3) |
|  | Other | 13 (0.92) | 27 (1.4) |
|  | Missing | 2 (0.28) | 0 (0) |
| Wave first started work (n (%)) |  |  |  |
|  | 8 (2016-2018) | 273 (19.3) | 303 (16.0) |
|  | 9 (2017-2019) | 288 (20.3) | 484 (25.6) |
|  | 10 (2018-2020) | 282 (20.0) | 337 (17.8) |
|  | 11 (2019-2021) | 202 (14.3) | 308 (16.3) |
|  | 12 (2020-2022) | 181 (12.8) | 227 (12.0) |
|  | 13 (2021-2023) | 186 (13.2) | 231 (12.2) |
| Education |  |  |  |
|  | No Degree | 885 (62.7) | 1097 (58.0) |
|  | Degree or higher | 510 (36.1) | 766 (40.5) |
|  | Missing | 17 (1.2) | 27 (1.4) |
| NS-SEC |  |  |  |
|  | Low (lower supervisory & technical, and semi-routine & routine) | 718 (50.9) | 983(52.0) |
|  | High (management & professional, intermediate, small employers & own account) | 482 (34.1) | 718 (38.0) |
|  | Missing | 212 (15.0) | 189 (10.0) |
| Parental Education |  |  |  |
|  | No Degree | 688 (48.7) | 823 (43.5) |
|  | Degree or higher | 635 (45.0) | 826 (43.7) |
|  | Missing | 89 (6.3) | 241 (12.8) |
| Job Characteristics |  |  |  |
| Commute Behaviour |  |  |  |
|  | Inactive | 551(39.0) | 914 (48.4) |
|  | Active | 508 (36.0) | 656 (34.7) |
|  | No Commute | 66 (4.7) | 62 (3.3) |
|  | Missing | 287 (20.3) | 258 (13.7) |
| Work Location |  |  |  |
|  | Office Work | 1073 (76.0) | 1590 (84.1) |
|  | Work from home | 66 (4.7) | 62 (3.3) |
|  | Driving around/ Travel Work | 137 (9.7) | 110 (5.8) |
|  | Missing Responses | 136 (9.6) | 128 (6.8) |
| Shift Times |  |  |  |
|  | Daytime | 527 (37.0) | 688 (36.4) |
|  | Night-time | 90 (6.4) | 101 (5.3) |
|  | Rotating | 308 (21.8) | 517 (27.4) |
|  | Missing | 487 (34.5) | 584 (30.9) |
| Work Hours |  |  |  |
|  | Full Time (30+ hours) | 841 (59.6) | 956 (50.6) |
| Part Time (less than 30) | 444 (31.4) | 830 (43.9) |
| Missing | 127 (9.0) | 104 (5.5) |

##### Individual characteristics (coded as time invariant)

**Sex** (“Male” [0], “Female” [1]); **Education (**collapsed into categories: “no degree”[0] or “at least a degree”[1]); **Parent’s Education status (**collapsed into categories: (“no degree”[0] or “at least a degree”[1]); **Ethnicity (**collapsed into categories: (“White”[0], “Asian” [1], “Black” [2], “Multiple” [3], “Other” [4]).

### Statistical analyses

#### Descriptive statistics

Analyses were performed using R Version 2023.12.0+369. Descriptive data on outcome and moderator variables were reported by sex.

Models were built up in line with interrupted time series regression, using the following formula:

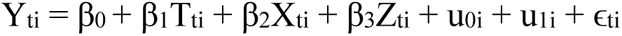

Multilevel linear models were fitted separately for each of the health behaviour outcomes (Y_ti_). Weights were not used due to the complexity of the model and due to the focus on within-person changes in outcomes. Models were built up sequentially, adding first the random intercept (u_0i_), then change in outcome over time in years (T_ti_); starting work (X_ti_) to assess an immediate effect of starting work, and time relative to starting work (Z_ti_), indicating time passed since starting work. The model assesses the sustained effect of entering the workforce and how this evolves over time, by allowing an additional intercept and a change in slope in response to starting work. Likelihood ratio tests were conducted to determine the best model fit for each outcome (*Supplementary Table 1*).

Data on fruit and vegetable consumption and sleep were roughly normally distributed, however physical activity data showed strong positive skew and heteroskedasticity of residuals. We considered applying various transformations but these limited interpretability and a log transformation would have implied an exponential growth curve, which was not a good fit with our data. We therefore decided to model the untransformed data using robust standard errors, benefiting from the robustness of multilevel models to violation of distributional assumptions (36).

We added moderator terms between time in years (T_ti_), starting work (X_ti_) and time relative to starting work (Z_ti_), to explore differences in intercept and slopes between different groups. The interaction with job characteristic models were adjusted for confounders including sex, ethnicity, parental education status, NS-SEC of their first job, and own education status. The interaction with education models were adjusted for sex, ethnicity, and parental education status. The interaction with ns-sec models were adjusted for education, sex, ethnicity, and parental education status (see *Supplementary Figure 2* for Directed Acyclic Graphs). Robust standard errors were used to generate 95% Confidence Intervals for all reported outcomes.

A sensitivity analysis using only participants who had exact job start dates from the wave they first started working was conducted to get a more accurate estimation of changes in behaviour before and after starting employment.

Graphs were generated using the “ggplot” and “predictions” packages in R to generate predicted values from the models to visualise changes in behaviour through the transition into employment. The full Rmarkdown html files for this analysis are available at: https://osf.io/wbscr/?view_only=fa9d385bb81e4c709fcb3fc11b0896e3

## Results

### Descriptive statistics

A total of 3,302 participants met the inclusion criteria and were included in the analyses (*Figure 1).* The mean age of participants when first starting working as their main occupation was 21.5 years (s.d.: 3.3), 57.2% were female, and 38.8% had a degree (*Table 1).* 184 people were included in a Supplementary Analysis where an exact job start date was reported *(Supplementary Table 1)*.

### Interrupted Time Series Analysis

The main results are reported in *Tables 4-7* and *Figure 2*. The unadjusted models are in the *Supplementary Tables 6-9* in the *Appendix* and were not dissimilar to the main results. Due to issues with singularity, random slope terms u_1i_ were not included in the vegetable intake models. Random slope terms were also not included in the physical activity or sleep models as they did not improve model fit, but were included in the fruit intake model. The addition of quadratic time variables was tested to allow the change in exposure over time to be non-linear. Addition of these quadratic terms did improve model fit, but the coefficients of these terms were very small and not significant, therefore they were dropped from the model to aid easy interpretation (*Supplementary table 1*).

**Figure 2.**
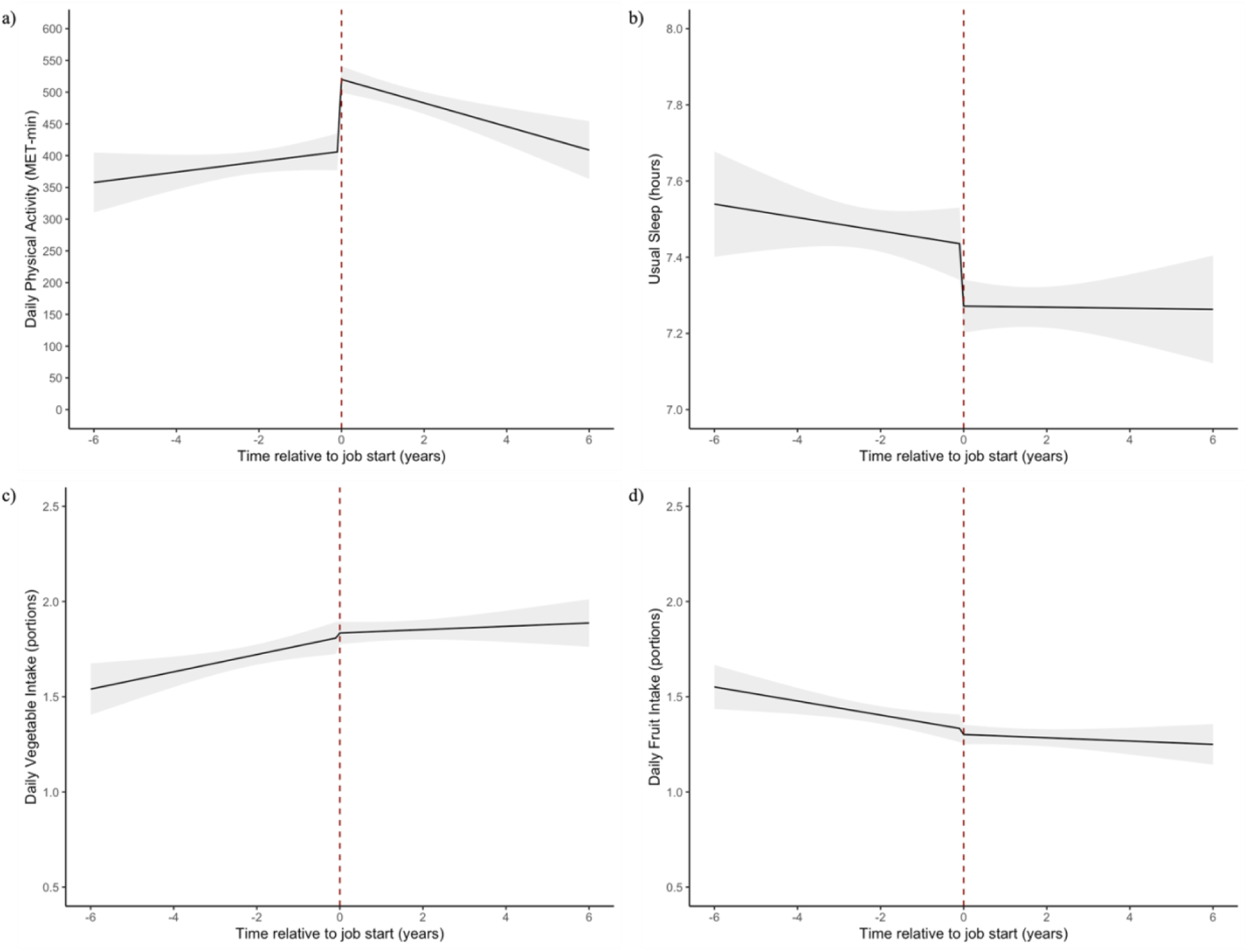
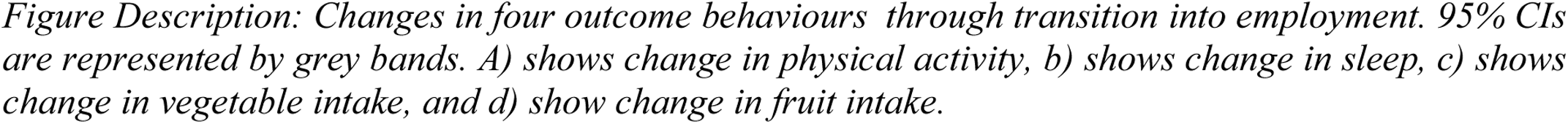
Predicted trajectories of physical activity, sleep, vegetable and fruit intake across the transition into employment.

**Figure 3:**
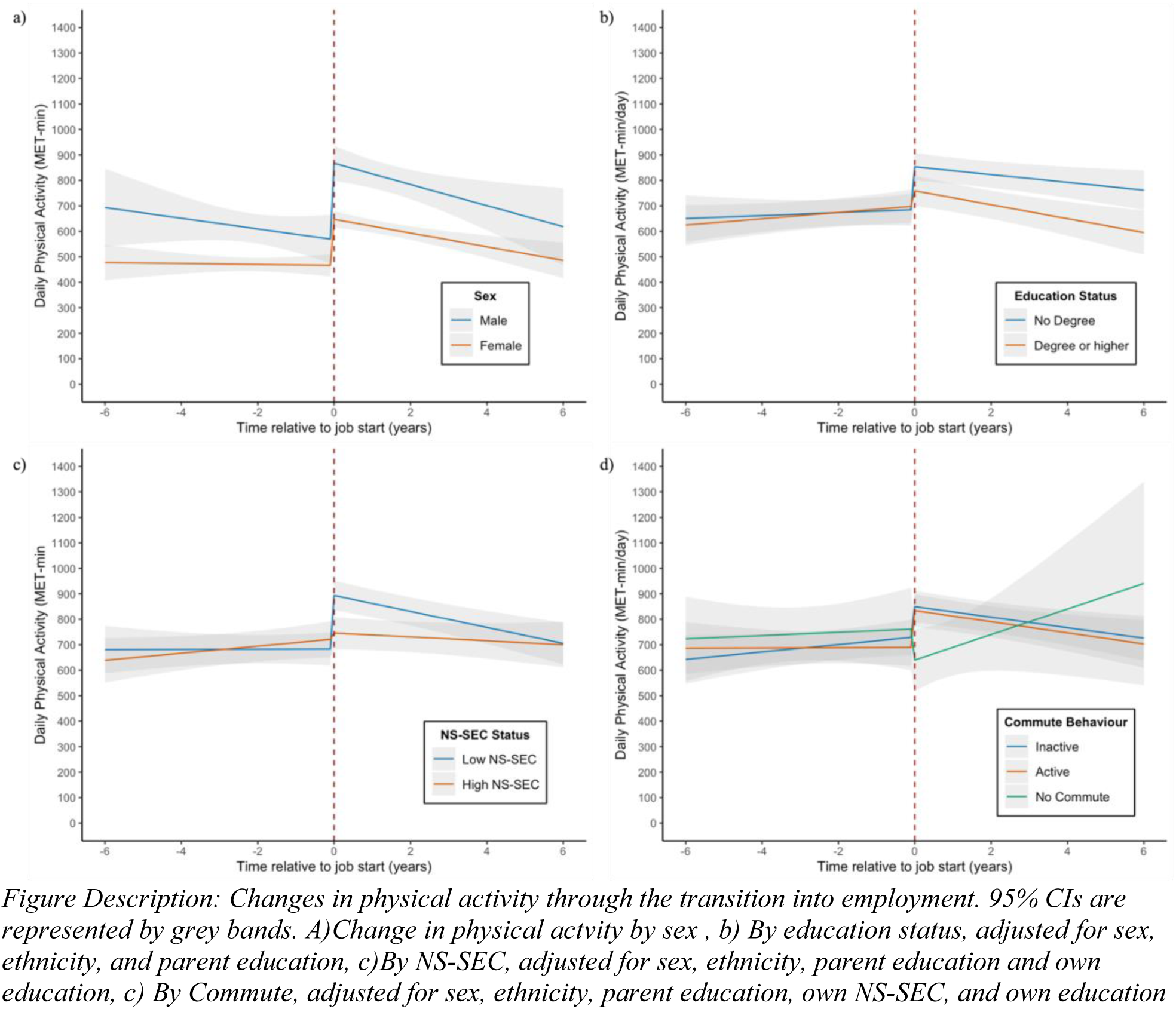
Physical activity (MET-min/day) trajectories across interaction groups

### Behavioural Trajectories through transition into employment

#### Changes in physical activity through the employment transition

Physical activity increased as people started work (*β=* 113.3, [95% CI: 80.49 to 146.11] MET-min/day) and then decreased over time after starting work (*β=* −26.7, [95% CI: −40.75 to - 12.66] MET-min/day/year) *(Figure 2, Table 4)*.

**Table 4:** MET-min per day over time.

|  |  | Initial slope [95% CI] | Transition into work [95% CI] | Change in slope after starting work [95% CI] |
| --- | --- | --- | --- | --- |
| Overall Effect<br>(n= 3,080) |  | 8.15 [-1.53,17.84] | 113.3 [80.49,146.11] | -26.7 [-40.75,-12.66] |
| Sex<br>(n= 3,080) | Male | -1.95 [-18.39,14.49] | 180.97 [124.39, 237.55] | -24.92 [-49.04,-0.8] |
|  | Female | 17.07 [5.97,28.17] | 62.58 [24.66,100.5] | -29.33 [-45.62,-13.04] |
|  | Interaction p-value | 0.06 | 0.001 | 0.766 |
| University Education<br>(n= 2,753) | Attended university | 12.46 [0.7,24.21] | 59.47 [14.91,104.02] | -39.19 [-57,-21.38] |
|  | No university | 6.9 [-11.3,25.09] | 164.02 [111.14,216.9] | -22.42 [-46.17,1.33] |
|  | Interaction p-value | 0.614 | 0.003 | 0.268 |
| NSSEC (n= 2,440) | High | 12.83 [-4.71, 30.37] | 13.95 [-44.35, 72.24] | -17.12 [-39.69, 5.44] |
|  | Low | 3.10 [-16.25, 22.46] | 204.17 [150.93, 257.42] | -32.88 [-55.80, -9.95] |
|  | Interaction p-value | 0.465 | <0.001 | 0.337 |
| Work Hours<br>(n= 2,397) | Full-time | -0.81 [-18.61,16.98] | 116.32 [61.78,170.86] | -16.85 [-40.47,6.77] |
|  | Part-time | -0.89 [-22.44,20.66] | 141.12 [80.73,201.5] | -13.57 [-41.87,14.74] |
|  | Interaction p-value | 0.995 | 0.547 | 0.861 |
| Shift Times (n=1,793) | Day time | -4.65 [-26.51, 17.21] | 128.49 [66.35, 190.62] | -9.78 [-35.58,16.02] |
|  | Night | -6.75 [-64.46, 50.96] | 204.47 [45.97, 362.98] | -35.23<br>[-102.02, 31.57] |
|  | Interaction p-value | 0.947 | 0.382 | 0.486 |
|  | Rotate | 14.80 [-11.65, 41.24] | 92.10 [16.72, 167.47] | -37.28 [-69.02, -5.55] |
|  | *Interaction p-value* | *0.265* | *0.465* | *0.187* |
| Commute (n= 2,183) | Inactive | 14.56 [-4.04, 33.16] | 119.56 [64.64, 174.48] | -35.19 [-57.96, -12.42] |
|  | Active | 0.51 [-20.15, 21.16 ] | 144.45 [83.60, 205.29] | -22.35 [-47.44, 2.74] |
|  | *Interaction p-value* | *0.320* | *0.551* | *0.457* |
|  | No Commute | 6.57 [-36.19, 49.34] | -122.29<br>[-297.33, 52.75] | 43.53 [-39.24,<br>126.30] |
|  | *Interaction p-value* | *0.737* | *0.010* | *0.072* |
| Work Location<br>(n= 2,356) | Office | 5.98 [-6.67,18.63] | 125.71 [85.79,165.63] | -25.27 [-43.23, -7.31] |
|  | Home | 15.53 [-20.48,51.54] | -157.81<br>[-313.07, -2.56] | 31.77 [-24.31,87.86] |
|  | Interaction p-value | 0.623 | 0.001 | 0.057 |
|  | Travel | 0.05 [-41.32,41.42] | 230.57 [86.84,374.31] | -45.36 [-104.43,13.7] |
|  | Interaction p-value | 0.788 | 0.167 | 0.523 |

Physical activity increased more in men (*β* =180.97 [95% CI: 124.39 to 237.55] MET-min/day) than in women (*β=* 62.58 [95% CI: 24.66 to 100.5] MET-min/day), (*Figure 2*). Participants who did not have a university degree showed a greater increase in physical activity (*β=* 164.02 [95% CI: 111.4 to 216.9] MET-min/day) compared to those with a university degree (*β=* 59.47 [95% CI: 14.91 to 104.02] MET-min/day). Working from home was associated with an initial decrease in physical activity (*β=* −157.81, [-313.07 to −2.56] MET-min/day), whereas those who worked in an office/employer’s premises showed an initial increase (*β=* 125.71 [95% CI: 85.79 to 165.63] MET-min/day), although this difference was not maintained.

### Changes in sleep behaviour through the employment transition

Usual minutes of sleep per night stayed stable over time (*β* = −1.05 [95% CI: −3.15 to 1.04] min/night/year), and then decreased immediately after starting work (*β* =-9.74 [95% CI: - 16.81 to −2.67], min/night). After starting work, sleep did not change over time (*β=* 0.97, [95% CI: −1.64 to 3.58] min/night/year) (*Figure 2, Supplementary Table 3*). Sleep behaviours did not differ by sex or NS-SEC status. Participants who did not have a university degree showed a decrease in sleep over time after starting work (*β* = −2.63 [95% CI: −7.36 to 2.10] min/night/year), whereas those with a university degree showed an increase over time (*β* = 3.58 [95% CI: −0.09 to 7.06] min/night/year) *(Figure 4b).* There were no differences in sleep behaviours based on job characteristics (*Supplementary Table 3*).

**Figure 4:**
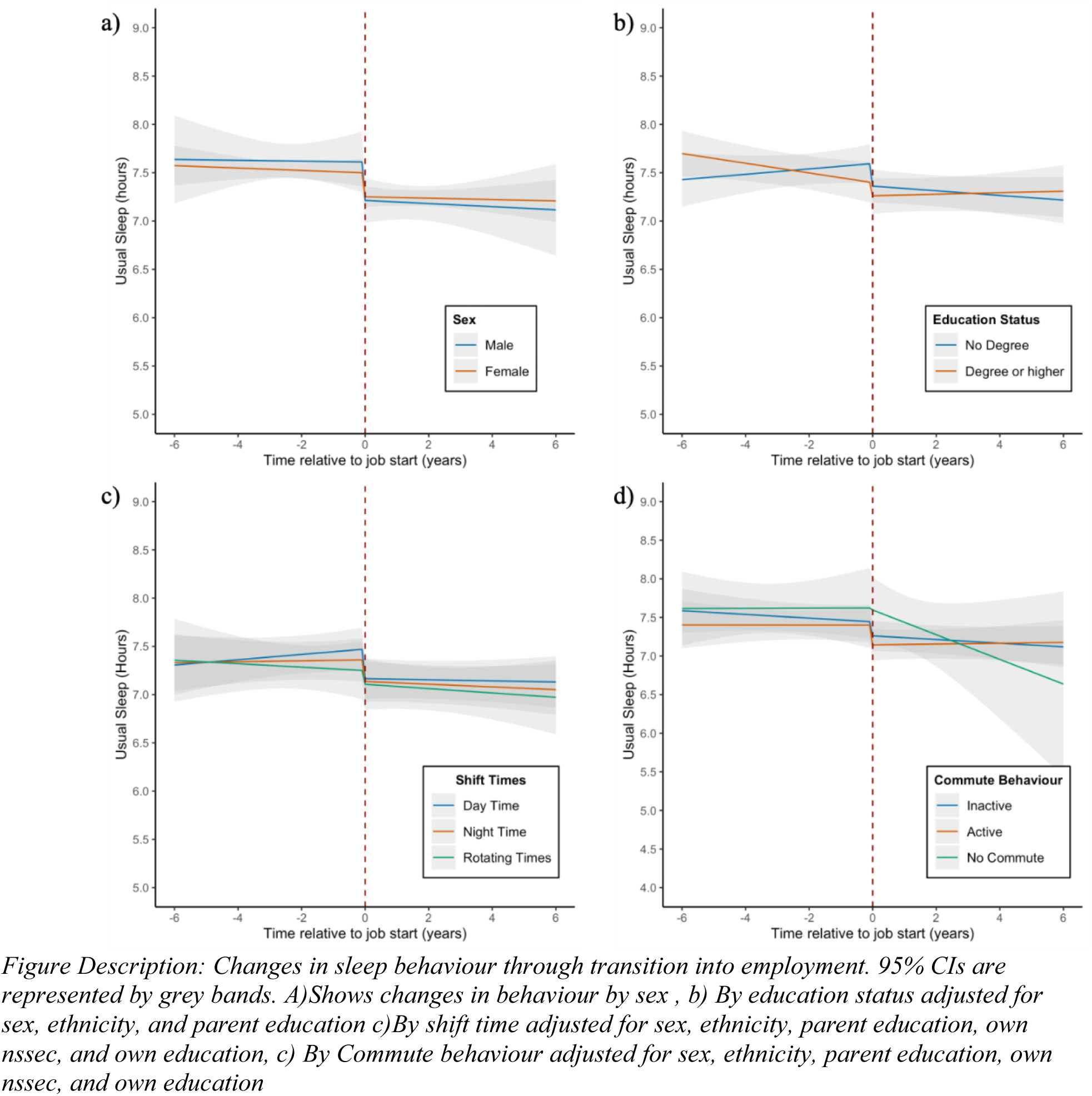
Sleep trajectories by individual and job characteristics

### Changes in diet behaviour through the employment transition

Daily vegetable portions showed a slight 0.05 portion/day increase over time [95% CI: 0.01 to 0.08] but starting work (*β*= 0.02 [95% CI: −0.07 to 0.12] vegetable portions/day) and time in employment (*β* = −0.40 % [95% CI: −0.80 to 0.00] vegetable portions/day/year) had no significant effect on vegetable intake (Figure 2, Supplementary *Table 4*). The transition into work was not associated with a significant change based on any individual or job characteristics.

Daily fruit portions showed a decrease of −0.04 portions/day each year before starting work [95% CI: −0.07 to −0.01 portions/day/year]. Starting work and time in employment had no significant effect on fruit intake (Figure 2, Supplementary *Table 5*). There were no differences in fruit intake based on sex. There was a significant interaction effect based on education status (*p=0.035*), such that participants who did not attend university showed a decrease in daily fruit intake after starting work (*β* = −0.12, [95%CI: −0.23 to 0.00] portions/day) and those who had a university degree showed an increase after starting work (*β* =0.06 [95% CI: −0.06 to 0.18] portions/day). No differences in fruit consumption were found based on job characteristics.

A supplementary analysis of changes in health behaviours through the employment transition using participants who reported an exact job start date (n=184) is reported in *Supplementary Table 2* in the Appendix. The results do not differ directionally from our main results.

## Discussion

Analysis of changes in physical activity, usual sleep, and diet suggest that starting work for the first time has important implications for developing health behaviours in young adults. Our findings showed that the greatest change in response to starting work was seen in physical activity, and moderate changes were seen in usual sleep behaviours. We found little association between starting work and changes in diet. Physical activity showed an initial increase on starting work of 113.3 MET-min/day, comparable to about 30 minutes of moderate activity per day (e.g. cycling at a regular pace) (32). The increase in physical activity was greatest among those of lower socioeconomic position, e.g. lower education and lower NS-SEC, as well as among men. Sleep patterns displayed a modest decline, on average 9.7 minutes, as young people entered the workforce, and then remained stable over the following years. After adjusting for confounders, only education status interacted with changes in sleep, such that people without a degree showed a continuing decline of about 2.6 usual minutes of sleep per year after starting work, while those with a degree showed a positive trajectory of usual minutes of sleep per night after starting work. Starting work showed limited associations on changes in diet. There was no overall effect of entering employment on vegetable intake, and no differences were seen based on any individual or job characteristics. No overall changes in fruit intake were seen after starting work, although those with no higher education showed a short-term decrease in daily intake after starting work compared to those with a university degree or higher.

### Implications of the findings and comparison with previous research

Increases in physical activity were more pronounced among individuals who are more likely to work in manual labour jobs, such as men and those with lower socioeconomic positions, likely due to their work requiring more physical activity (37). The differences in physical activity due to education status persisted over time, with people with a university degree consistently reporting lower levels of activity than those with no degree after starting work. Although stratifying by participants’ NS-SEC status showed similar differences immediately after starting work, these differences did not remain over time. People with a low NS-SEC job showed a steeper decline in physical activity each year after starting work, such that the difference in activity levels between those low NS-SEC and high NS-SEC groups grew smaller over time. This suggests that education status and NS-SEC have separate mechanisms on physical activity levels. The NS-SEC effect may be related to job role, which may well change over time as people get more senior.

Although physical activity increased more after starting employment among groups more likely to encounter physically demanding work (37–39), a previous meta-analysis found that occupational physical activity may not have the same benefits in terms of cardiovascular outcomes compared to leisure-time physical activity (40). More research is needed to differentiate between the longitudinal effects of occupational physical activity and ‘leisure-time’ physical activity, as there may be negative long-term effects of work-related stressors and work quality related to occupational physical activity.

Commuting behaviours also seemed to play a role in changes in physical activity through the transition of starting work. Those with no commute showed a nonsignificant decrease in physical activity after starting work compared to increases in physical activity in both active and inactive commuters. Relatedly, working from home also showed decrease in physical activity compared to those who worked in offices or worked elsewhere out of the home. Previous studies have found that active commuting is associated with positive outcomes in physical activity and physical fitness compared to no active commutes (41,42).

While usual hours of sleep only showed a slight decrease after starting work, people without a university degree seemed to show more long-term negative effects after starting work. This is consistent with previous findings from the US Behavioral Risk Factor Surveillance study that found associations between low occupational social class and sleep disturbances (43). While our study did not find any associations between shift work and sleep behaviour, a systematic review reported that both psychosocial and shift times are associated with sleep disturbances (22). It may be that sleep quality is more strongly influenced by the workplace than sleep duration. Sleep and work have previously been connected in a negative cycle where poor sleep quality is associated with more negative events and perceived stress the following day, and these negative events are also associated with poorer sleep quality the following night (44–46).

We found very limited changes in fruit and vegetable intakes after starting work, which does not align with previous work on workplace environments and diet, as well as other longitudinal studies assessing life transitions and diet (18,19,28,32,47). However, these previous studies were all conducted in different contexts (mainly the U.S. and Norway).

### Strengths and limitations

This the first study to examine how health behaviours change in young adults when they start employment in the UK. The study design using a multilevel interrupted-time series analysis improves causal inference compared to other study designs by including a baseline trend before starting work and allowing modelling of both immediate and long-term effects of starting work.

We use data from a national panel survey, representative of the UK population, increasing the generalizability of the findings within the UK context. The survey included detailed questions on job characteristics as well as health behaviour measures, over many years, allowing for longitudinal analysis. However, all the variables used were self-reported, as there was no objective measurement of physical activity, sleep, or diet in this dataset, providing opportunity for reporting bias. Additionally, the diet variables only assessed fruit and vegetable intake, and while increased fruit and vegetable consumption have been linked to a better overall diet (48), they do not provide a comprehensive insight into overall diet quality. Given the limited availability of longitudinal surveys that provide high-quality, repeated measures of diet quality over the early adulthood period, the available fruit and vegetable intake variables were considered appropriate for this analysis.

Our study included a large sample size of over 3,000 participants, providing data across multiple time points. Despite this, some groups (e.g., work-from-home or night-time and rotating shift workers) had small numbers, which may contribute to small effect sizes and large confidence intervals. Additionally, due to missing data regarding exact job start dates, the interview date from participants’ first wave of reported working was used instead, which may limit the accuracy and interpretation of results. We may not see the true immediate effect, as participants could have up to 2 years from when the transition takes place to their first recorded data point. However, a sensitivity analysis that only included participants with an exact job start date showed similar direction and magnitude of findings.

## Conclusions

This analysis revealed a clear impact of starting work on changes in physical activity and sleep among young adults. A positive impact of starting work was seen on physical activity, and a a negative impact through reduction in sleep duration. Changes in physical activity in response to starting work differed by job characteristics, highlighting the potential influence of the work environment on health behaviours. This suggests that the workplace could be a good opportunity for health promotion. Workplace health promotion interventions promoting healthier diets, physical activity, and sleep in young adults could result in healthier employees and fewer sick days. Targeting young adulthood for health behaviour interventions could prevent future health issues, combat rising obesity rates, and promote positive generational changes. This study highlights the importance of understanding how employment transitions impact young adults’ health behaviours and the potential for policy interventions to support healthier lifestyles among young working adults. Future research should focus on the interplay between individual characteristics, job attributes, and job quality in shaping long-term health outcomes.

## Data Availability

The data that support the findings of this study are available on request from the UK Data Service, study number (SN) 6614. The data are publicly available, however, they are considered safeguarded and therefore require users to register and accept the End User Licence.

https://www.understandingsociety.ac.uk/documentation/access-data/

## Declarations

### Ethics approval and consent to participate

The University of Essex Ethics Committee has approved all data collection on Understanding Society main study, COVID-19 surveys and innovation panel waves, including asking consent for all data linkages except to health records.

### Consent for publication

Not Applicable

### Availability of data and materials

The data that support the findings of this study are available on request from the UK Data Service https://doi.org/10.5255/UKDA-SN-6614-19, study number (SN) 6614. The data are publicly available, however, they are considered safeguarded and therefore require users to register and accept the End User Licence.

All Rmarkdown files are available on OSF at: https://osf.io/wbscr/?view_only=fa9d385bb81e4c709fcb3fc11b0896e3

### Authors’ contributions

AFO conducted the analysis and drafted the manuscript. EMW, AM, EvS and HF supported the conceptualization of the study, interpreting results and drafting, reviewing and editing the manuscript. TB provided additional guidance on coding in R. All authors reviewed drafts of the manuscript.

## Acknowledgements

We would like to acknowledge the contributions of Dr. Yinhua Tao for guidance on statistical analysis and visualisation methods, as well as thank the study participants for their contribution in providing the data used in this analysis. We would also like to thank the members of our Young Persons Advisory Group and project stakeholders for their feedback.

Understanding Society is an initiative funded by the Economic and Social Research Council and various Government Departments, with scientific leadership by the Institute for Social and Economic Research, University of Essex, and survey delivery by the National Centre for Social Research (NatCen) and Verian (formerly Kantar Public). The research data are distributed by the UK Data Service. The COVID-19 study (2020–2021) was funded by the Economic and Social Research Council and the Health Foundation. Serology testing was funded by the COVID-19 Longitudinal Health and Wealth – National Core Study. Fieldwork for the web survey was carried out by Ipsos MORI and for the telephone survey by Kantar.

## Appendix

**Supplementary Table 1:**
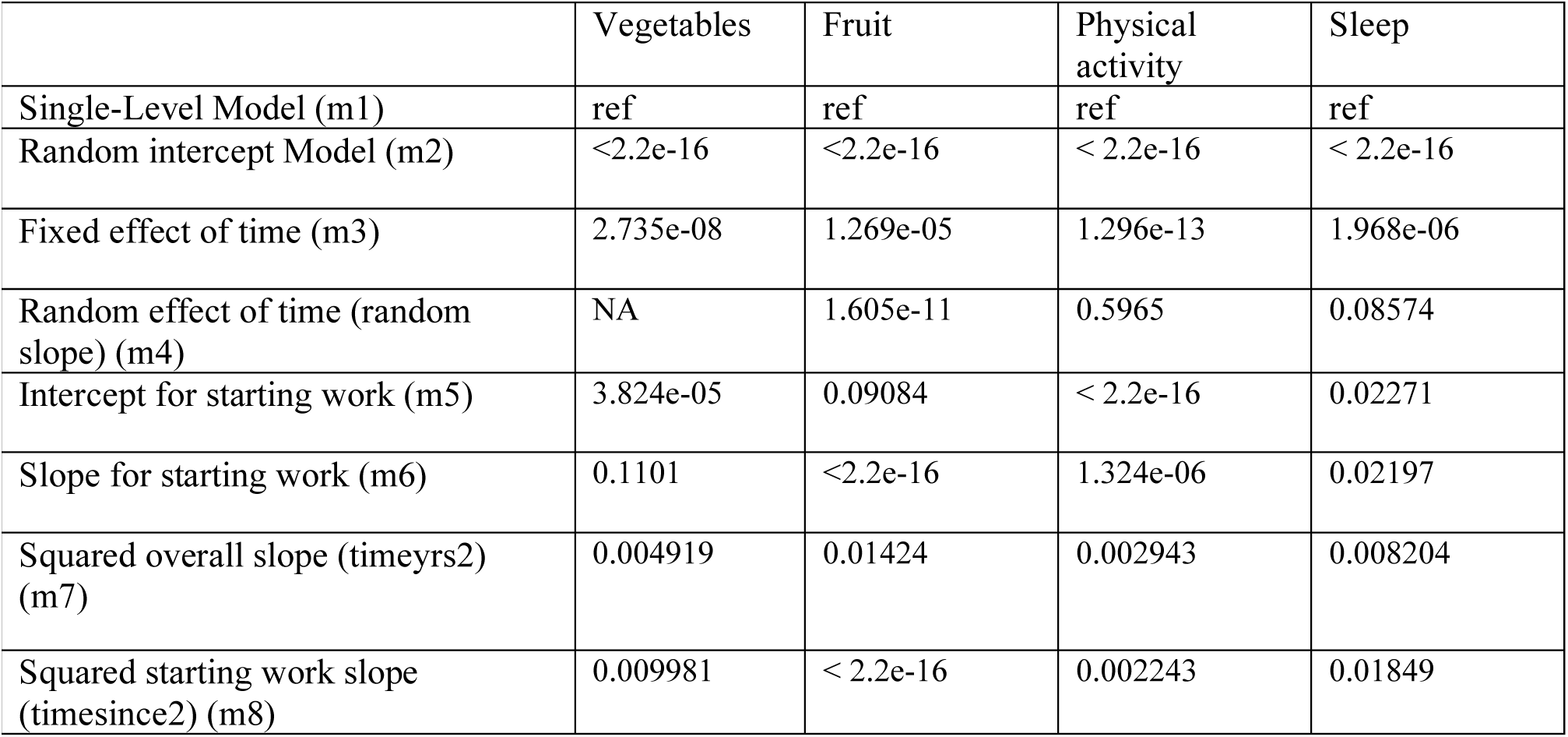
Likelihood ratio test for comparison with previous model.

**Supplementary Table 2:**
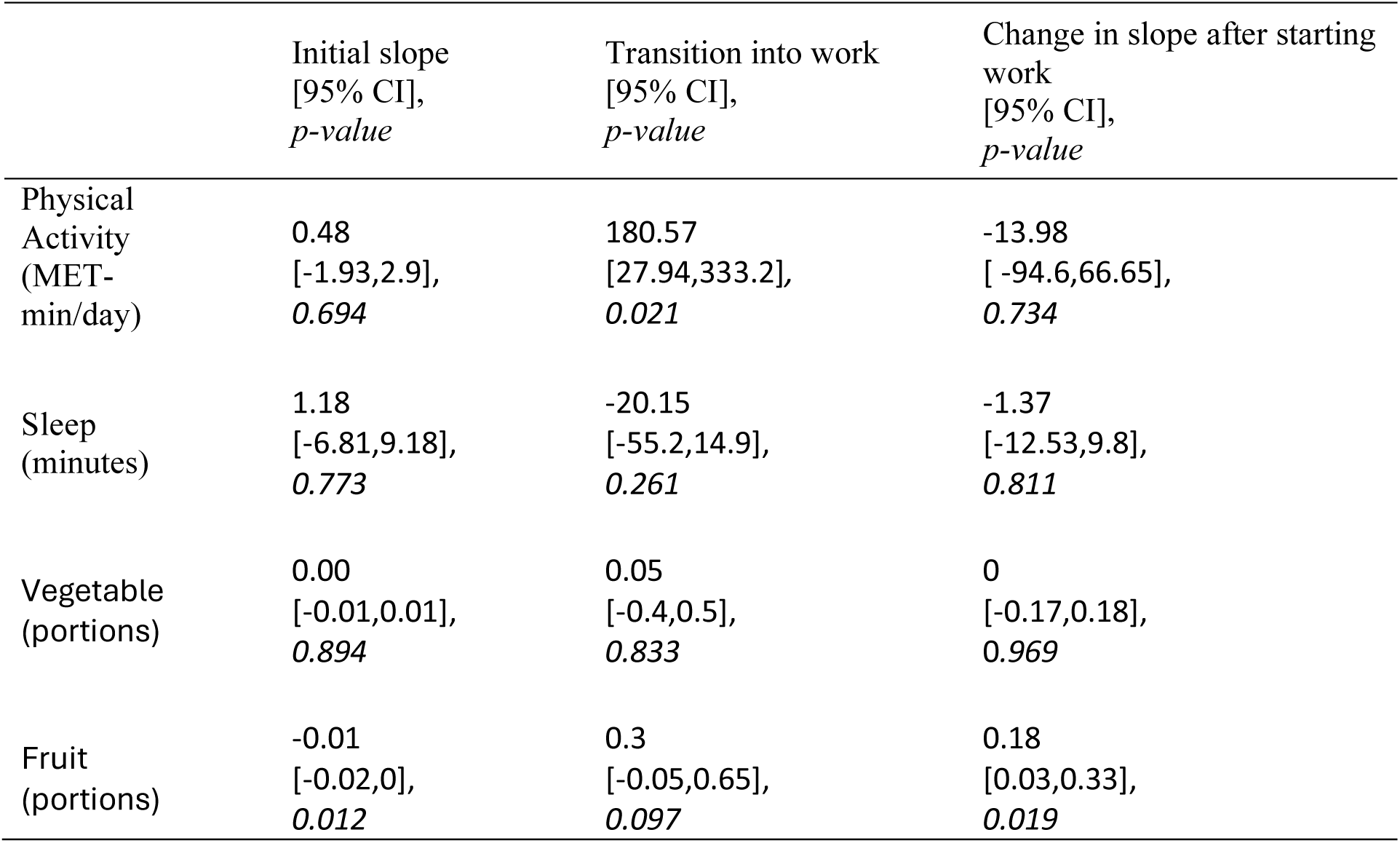
Supplementary analysis of four health behaviours, only including participants with an exact job start date.

|  | Initial slope<br>[95% CI],<br>*p-value* | Transition into work<br>[95% CI],<br>*p-value* | Change in slope after starting<br>work<br>[95% CI],<br>*p-value* |
| --- | --- | --- | --- |
| Physical Activity<br>(MET-min/day) | 0.48<br>[-1.93,2.9],<br>*0.694* | 180.57<br>[27.94,333.2],<br>*0.021* | -13.98<br>[ -94.6,66.65],<br>*0.734* |
| Sleep<br>(minutes) | 1.18<br>[-6.81,9.18],<br>*0.773* | -20.15<br>[-55.2,14.9],<br>*0.261* | -1.37<br>[-12.53,9.8],<br>*0.811* |
| Vegetable<br>(portions) | 0.00<br>[-0.01,0.01],<br>*0.894* | 0.05<br>[-0.4,0.5],<br>*0.833* | 0<br>[-0.17,0.18],<br>*0.969* |
| Fruit<br>(portions) | -0.01<br>[-0.02,0],<br>*0.012* | 0.3<br>[-0.05,0.65],<br>*0.097* | 0.18<br>[0.03,0.33],<br>*0.019* |

**Supplementary Figure 1:**
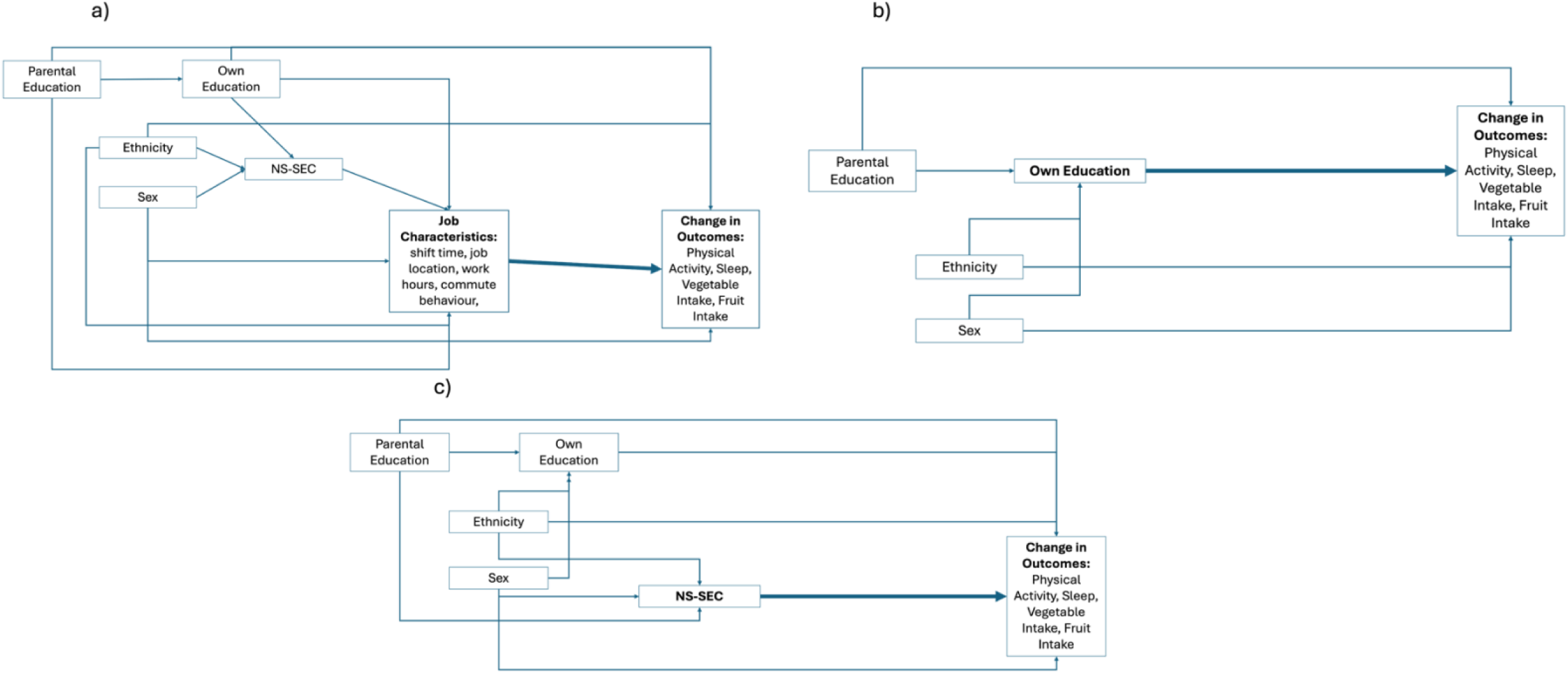
Directed Acyclic Graphs of interaction models and confounders

**Supplementary Table 3:**
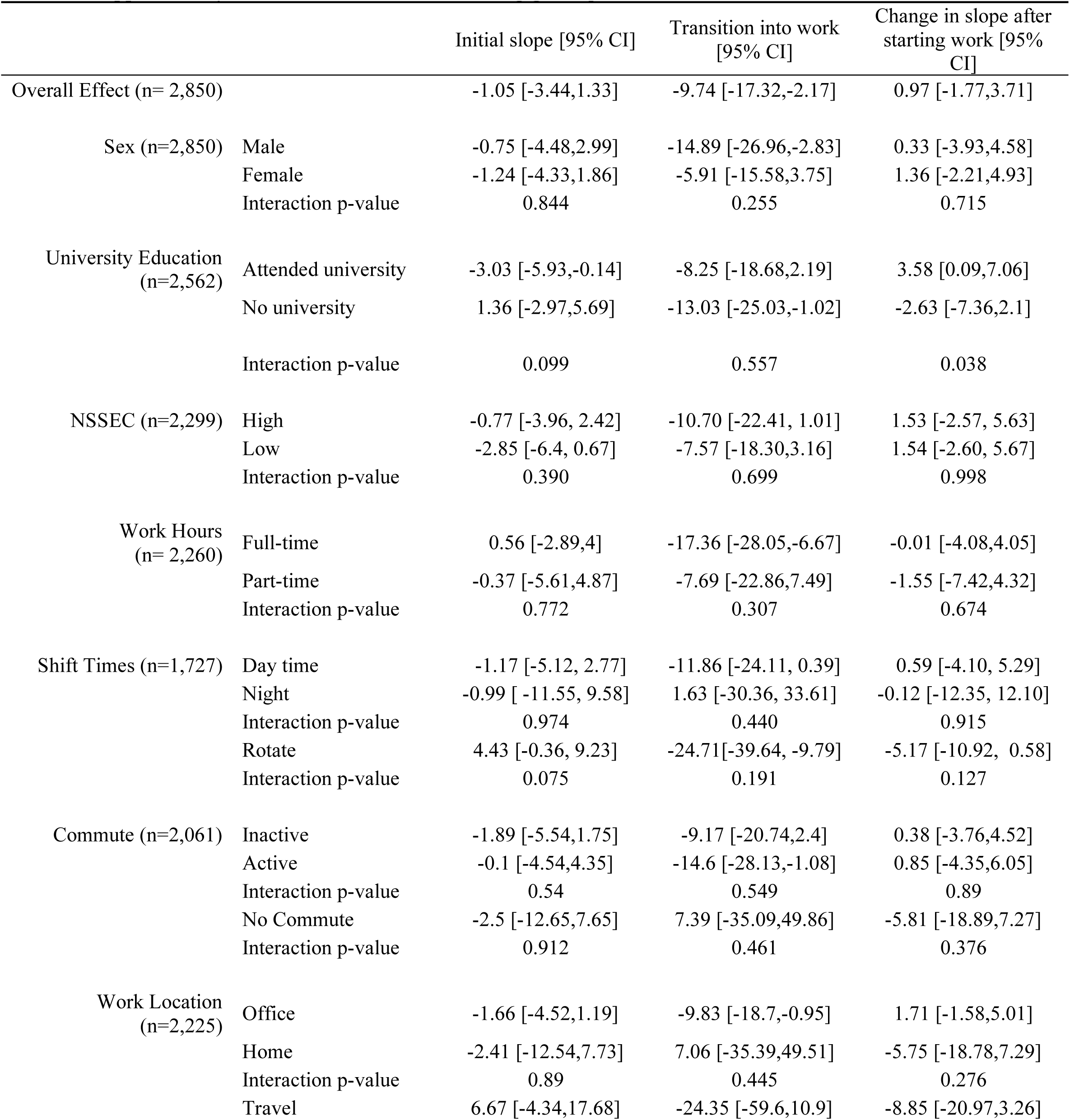
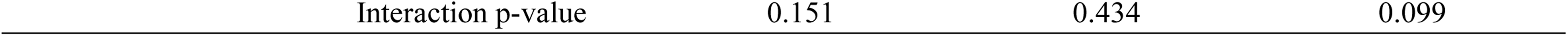
Usual Minutes of Sleep per night.

**Supplementary Table 4:**
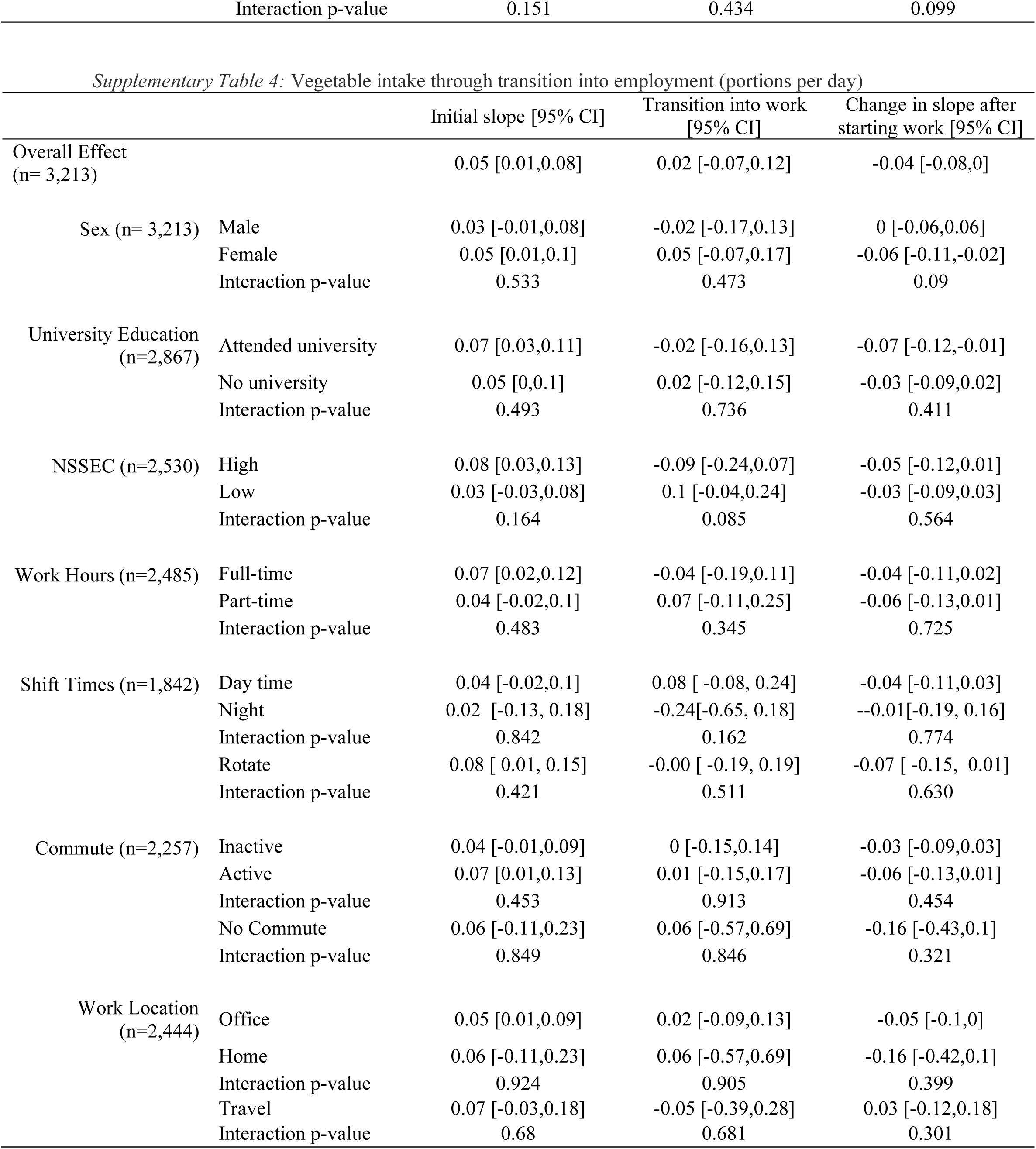
Vegetable intake through transition into employment (portions per day)

**Supplementary Table 5:** Daily portions of fruit.

|  |  | Initial slope [95% CI] | Transition into work<br>[95% CI] | Change in slope after<br>starting work [95% CI] |
| --- | --- | --- | --- | --- |
| Overall Effect<br>(n= 3,218) |  | -0.04 [-0.07,-0.01] | -0.03 [-0.11,0.05] | 0.03 [-0.01,0.06] |
| Sex (n= 3,218) | Male | -0.02 [-0.06,0.02] | -0.07 [-0.19,0.04] | 0 [-0.06,0.06] |
|  | Female | -0.05 [-0.09,0] | 0 [-0.1,0.11] | 0.02 [-0.03,0.07] |
|  | *Interaction p-value* | *0.477* | *0.343* | *0.694* |
| University Education<br>(n= 2,872) | Attended university | -0.03 [-0.07,0] | 0.06 [-0.06, 0.18] | -0.02 [-0.07,0.03] |
|  | No university | -0.02 [-0.07,0.03] | -0.12 [-0.23, 0] | 0.03 [-0.02,0.09] |
|  | *Interaction p-value* | *0.629* | *0.035* | *0.159* |
| NSSEC (n=2,534) | High | -0.03 [-0.08,0.01] | 0.01 [-0.13,0.14] | 0.01 [-0.05,0.06] |
|  | Low | -0.02 [-0.07,0.02] | -0.07 [-0.19,0.06] | 0.03 [-0.03,0.08] |
|  | *Interaction p-value* | *0.749* | *0.457* | *0.592* |
| Work Hours (n=2489) | Full-time | -0.03 [-0.08,0.03] | -0.02 [-0.15,0.11] | 0.02 [-0.04,0.08] |
|  | Part-time | -0.03 [-0.1,0.03] | -0.05 [-0.21,0.1] | 0.01 [-0.06,0.08] |
|  | *Interaction p-value* | *0.918* | *0.735* | *0.899* |
| Shift Times (n=1844) | Day time | -0.02 [-0.07 , 0.03] | -0.01 [-0.15, 0.14] | -0.00 [-0.06, 0.06] |
|  | Nighttime | -0.01 [-0.15, 0.13] | -0.17 [-0.53, 0.19] | 0.01 [-0.14, 0.17] |
|  | *Interaction p-value* | *0.884* | *0.416* | *0.850* |
|  | Rotating times | -0.02 [-0.09, 0.04] | -0.07 [-0.24, 0.10] | 0.00 [-0.07, 0.07] |
|  | *Interaction p-value* | *0.905* | *0.608* | *0.929* |
| Commute (n=2262) | Inactive | -0.03 [-0.08,0.02] | -0.05 [-0.17,0.08] | 0.03 [-0.03,0.09] |
|  | Active | -0.01 [-0.06,0.04] | -0.08 [-0.22,0.07] | -0.01 [-0.06,0.05] |
|  | *Interaction p-value* | *0.579* | *0.75* | *0.418* |
|  | No Commute | -0.05 [-0.21,0.11] | 0.17 [-0.42,0.75] | -0.08 [-0.33,0.16] |
|  | *Interaction p-value* | *0.832* | *0.482* | *0.378* |
| Work Location<br>(n=2,448) | Office | -0.02 [-0.06,0.01] | -0.07 [-0.16,0.03] | 0.02 [-0.03,0.06] |
|  | Home | -0.07 [-0.23,0.1] | 0.22 [-0.37,0.81] | -0.07 [-0.32,0.18] |
|  | *Interaction p-value* | *0.611* | *0.351* | *0.496* |
|  | Travel | -0.07 [-0.21,0.08] | 0.19 [-0.16,0.54] | 0.01 [-0.15,0.17] |
|  | *Interaction p-value* | *0.567* | *0.167* | *0.95* |

### Unadjusted Models

**Supplementary Table 6:**
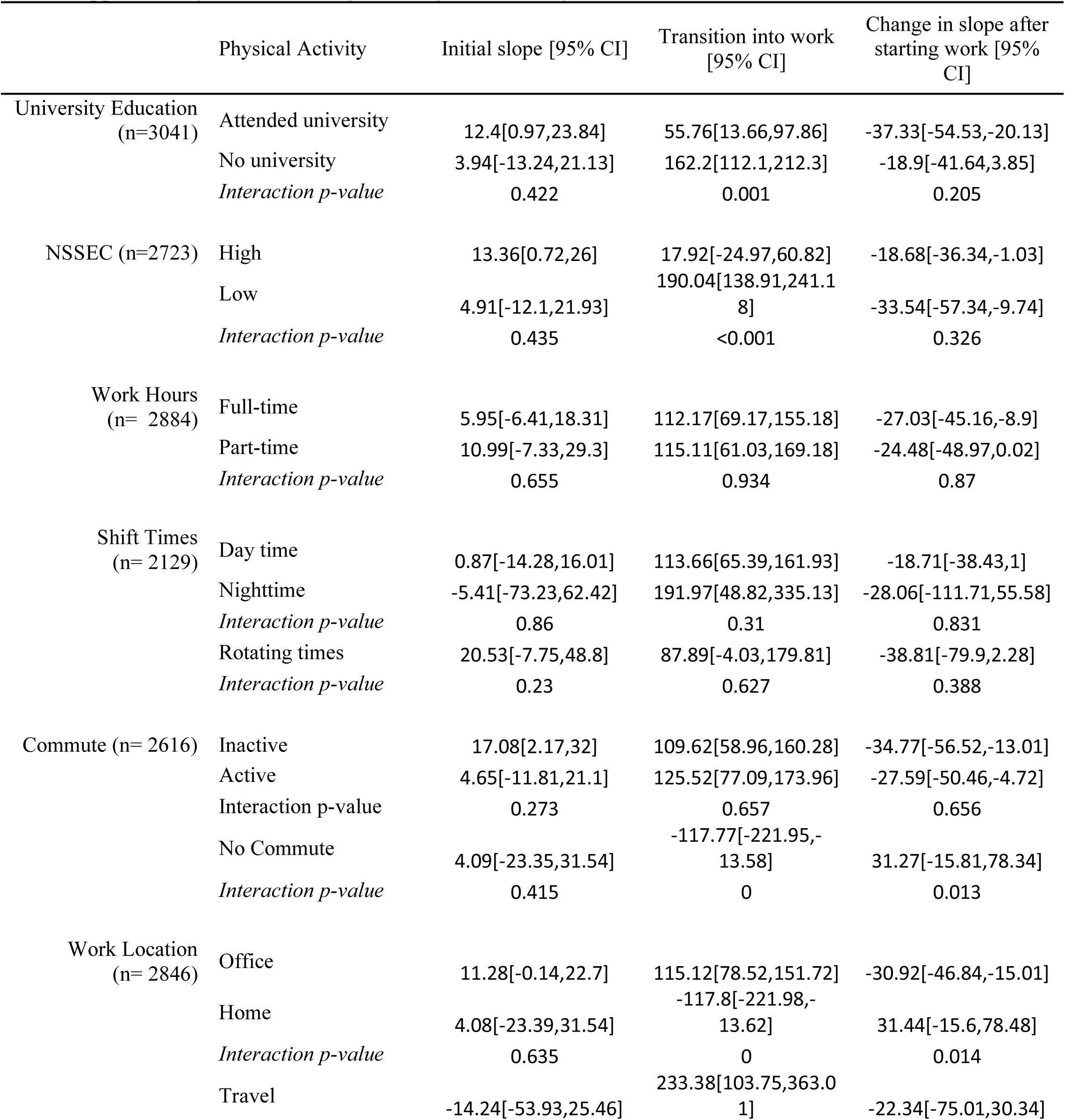
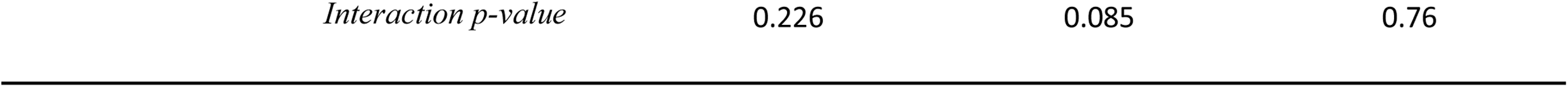
Unadjusted Physical Activity Models.

**Supplementary Table 7:** Unadjusted Sleep Models.

|  | Sleep | Initial slope [95% CI] | Transition into work [95% CI] | Change in slope after starting work [95% CI] |
| --- | --- | --- | --- | --- |
| University Education<br>(n=2819) | Attended university | -3.3[-6.11,-0.48] | -6.96[-16.93,3.02] | 4.41[1.04,7.77] |
|  | No university | 3.16[-1.15,7.46] | -16.2[-27.9,-4.5] | -4.26[-8.89,0.37] |
|  | *Interaction p-value* | 0.014 | 0.239 | 0.003 |
| NSSEC (n=2549) | High | 0.01[-3.33,3.35] | -10.8[-21.4,-0.19] | 0.61[-3.12,4.34] |
|  | Low | -2.11[-6.1,1.88] | -8.45[-20.13,3.23] | 1.62[-2.89,6.13] |
|  | *Interaction p-value* | 0.426 | 0.771 | 0.735 |
| Work Hours<br>(n= 2688) | Full-time | -0.59[-3.4,2.22] | -12.79[-21.88,-3.69] | 1.16[-2.1,4.42] |
|  | Part-time | -0.77[-5.68,4.14] | -7.65[-21.8,6.51] | -0.07[-5.45,5.3] |
|  | *Interaction p-value* | 0.951 | 0.549 | 0.701 |
| Shift Times<br>(n= 2036) | Day time | 1.98[-1.93,5.9] | -18.5[-29.63,-7.37] | -1.44[-5.73,2.86] |
|  | Nighttime | 1.6[-11.19,14.4] | -8.37[-42.83,26.1] | -1.72[-15.85,12.41] |
|  | *Interaction p-value* | 0.955 | 0.583 | 0.97 |
|  | Rotating times | -0.1[-5.22,5.02] | -9.72[-26.71,7.26] | -0.49[-6.92,5.94] |
|  | *Interaction p-value* | 0.525 | 0.397 | 0.81 |
| Commute (n=2447) | Inactive | -1.09[-4.52,2.34] | -11.78[-22.58,-0.97] | 0.53[-3.35,4.41] |
|  | Active | -0.81[-4.87,3.24] | -10.52[-22.73,1.7] | 1.81[-2.91,6.53] |
|  | *Interaction p-value* | 0.918 | 0.88 | 0.682 |
|  | No Commute | 1.35[-6.61,9.31] | -14.76[-48.18,18.66] | -6.51[-18.4,5.37] |
|  | *Interaction p-value* | 0.581 | 0.868 | 0.27 |
| Work Location<br>(n= 2658) | Office | -1.08[-3.82,1.66] | -8.77[-17.09,-0.44] | 1.25[-1.85,4.34] |
|  | Home | 1.44[-6.5,9.38] | -14.97[-48.39,18.45] | -6.52[-18.37,5.32] |
|  | *Interaction p-value* | 0.557 | 0.724 | 0.214 |
|  | Travel | 7.99[-1.97,17.95] | -38.11[-70.04,-6.18] | -7.65[-18.29,2.98] |
|  | *Interaction p-value* | 0.085 | 0.081 | 0.115 |

**Supplementary Table 8:** Unadjusted Vegetable Models.

|  |  | Initial slope [95%<br>CI] | Transition into<br>work [95% CI] | Change in slope<br>after starting work<br>[95% CI] |
| --- | --- | --- | --- | --- |
| University Education (n=3171) | Attended university | 0.06[0.01,0.1] | 0[-0.14,0.14] | -0.06[-0.11,0] |
|  | No university | 0.05[0,0.1] | 0.02[-0.11,0.15] | -0.03[-0.09,0.02] |
|  | *Interaction p-value* | 0.851 | 0.854 | 0.55 |
| NSSEC (n= 2830) | High | 0.06[0.01,0.11] | -0.02[-0.16,0.13] | -0.06[-0.12,0] |
|  | Low | 0.02[-0.03,0.06] | 0.12[-0.01,0.26] | -0.02[-0.08,0.04] |
|  | *Interaction p-value* | 0.179 | 0.168 | 0.299 |
| Work Hours (n=3000) | Full-time | 0.04[0,0.08] | 0.03[-0.1,0.16] | -0.04[-0.09,0.01] |
|  | Part-time | 0.04[-0.01,0.09] | 0.08[-0.07,0.23] | -0.04[-0.1,0.02] |
|  | *Interaction p-value* | 0.999 | 0.628 | 0.953 |
| Shift Times (n=2192) | Day time | 0.03[-0.02,0.08] | 0.08[-0.05,0.21] | -0.02[-0.08,0.04] |
|  | Nighttime | 0.04[-0.11,0.18] | -0.23[-0.63,0.16] | 0[-0.16,0.17] |
|  | *Interaction p-value* | 0.951 | 0.141 | 0.825 |
|  | Rotating times | 0.06[-0.01,0.14] | 0.14[-0.1,0.38] | -0.1[-0.19,-0.01] |
|  | *Interaction p-value* | 0.441 | 0.684 | 0.137 |
| Commute (n=2717) | Inactive | 0.03[-0.01,0.08] | 0.01[-0.12,0.14] | -0.02[-0.08,0.04] |
|  | Active | 0.05[0,0.1] | 0.06[-0.09,0.2] | -0.05[-0.11,0.02] |
|  | *Interaction p-value* | 0.623 | 0.658 | 0.556 |
|  | No Commute | 0.02[-0.13,0.17] | 0.2[-0.33,0.73] | -0.14[-0.37,0.1] |
|  | *Interaction p-value* | 0.849 | 0.502 | 0.345 |
| Work Location (n=2963) | Office | 0.04[0.01,0.08] | 0.05[-0.06,0.15] | -0.04[-0.09,0] |
|  | Home | 0.02[-0.13,0.17] | 0.2[-0.33,0.73] | -0.14[-0.37,0.1] |
|  | *Interaction p-value* | 0.76 | 0.582 | 0.445 |
|  | Travel | 0.04[-0.06,0.14] | 0.01[-0.29,0.3] | 0.06[-0.07,0.19] |
|  | *Interaction p-value* | 0.964 | 0.812 | 0.142 |

**Supplementary Table 9:** Unadjusted Fruit Models.

|  | FRUIT | Initial slope [95% CI] | Transition into work<br>[95% CI] | Change in slope after<br>starting work [95%<br>CI] |
| --- | --- | --- | --- | --- |
| University Education<br>(n=3177) | Attended university | -0.05[-0.08,-0.01] | 0.07[-0.05,0.18] | 0[-0.05,0.05] |
|  | No university | -0.02[-0.07,0.03] | -0.11[-0.22,0] | 0.04[-0.02,0.09] |
|  | *Interaction p-value* | 0.391 | 0.028 | 0.301 |
| NSSEC (n=2834) | High | -0.04[-0.08,0.01] | 0.02[-0.1,0.14] | 0[-0.05,0.06] |
|  | Low | -0.04[-0.09,0] | -0.03[-0.15,0.08] | 0.05[-0.01,0.1] |
|  | *Interaction p-value* | 0.898 | 0.543 | 0.262 |
| Work Hours<br>(n=3004) | Full-time | -0.04[-0.08,0] | 0.01[-0.1,0.11] | 0.03[-0.02,0.08] |
|  | Part-time | -0.05[-0.1,0.01] | -0.06[-0.19,0.07] | 0.04[-0.02,0.1] |
|  | *Interaction p-value* | 0.875 | 0.439 | 0.77 |
| Shift Times (n=2196) | Day time | -0.02[-0.07,0.02] | 0.01[-0.1,0.12] | 0[-0.05,0.05] |
|  | Nighttime | 0.01[-0.11,0.12] | -0.16[-0.5,0.17] | 0[-0.15,0.15] |
|  | *Interaction p-value* | 0.625 | 0.335 | 0.966 |
|  | Rotating times | -0.06[-0.14,0.03] | -0.1[-0.32,0.12] | 0.05[-0.04,0.15] |
|  | *Interaction p-value* | 0.472 | 0.392 | 0.338 |
| Commute (n=2724) | Inactive | -0.04[-0.08,0.01] | -0.02[-0.13,0.1] | 0.03[-0.02,0.09] |
|  | Active | -0.04[-0.09,0.01] | -0.05[-0.18,0.09] | 0.02[-0.03,0.08] |
|  | *Interaction p-value* | 0.923 | 0.739 | 0.773 |
|  | No Commute | -0.03[-0.15,0.08] | 0.1[-0.32,0.51] | -0.06[-0.26,0.13] |
|  | *Interaction p-value* | 0.944 | 0.612 | 0.33 |
| Work Location<br>(2969) | Office | -0.04[-0.07,0] | -0.04[-0.12,0.05] | 0.03[-0.01,0.07] |
|  | Home | -0.03[-0.15,0.08] | 0.1[-0.32,0.51] | -0.06[-0.26,0.13] |
|  | *Interaction p-value* | 0.953 | 0.538 | 0.331 |
|  | Travel | -0.09[-0.22,0.04] | 0.14[-0.17,0.44] | 0.05[-0.09,0.19] |
|  | *Interaction p-value* | 0.417 | 0.281 | 0.833 |

